# Early pregnancy health behaviors, adverse pregnancy outcomes, and maternal blood pressure after pregnancy

**DOI:** 10.1101/2024.12.27.24319672

**Authors:** Andrea C Kozai, Bethany Barone Gibbs, Lisa D Levine, Abbi D Lane, Mitali Ray, William Grobman, Philip Greenland, Matthew K Hoffman, C Noel Bairey Merz, Phyllis C Zee, Lisa Mims, Natalie A Cameron, George Saade, Robert M Silver, Natalie A Bello, Eliza C Miller, Uma M Reddy, Judith H Chung, Kara M Whitaker, Janet M Catov

**Affiliations:** Department of Epidemiology, University of Pittsburgh; Department of Epidemiology & Biostatistics, West Virginia University School of Public Health; Department of Obstetrics & Gynecology, University of Pennsylvania Perelman School of Medicine; Department of Applied Exercise Science, University of Michigan; Department of Health Promotion & Development, University of Pittsburgh School of Nursing; Department of Obstetrics & Gynecology, Brown University School of Medicine; Departments of Medicine (Cardiology) and Preventive Medicine, Northwestern University Feinberg School of Medicine; Department of Obstetrics & Gynecology, ChristianaCare; Department of Cardiology, Smidt Heart Institute, Cedars Sinai Medical Center; Department of Neurology, Northwestern University Feinberg School of Medicine; Department of Obstetrics & Gynecology, Indiana University School of Medicine; Departments of General Internal Medicine and Preventive Medicine, Northwestern University Feinberg School of Medicine; Department of Obstetrics & Gynecology, Eastern Virginia Medical School; Department of Obstetrics & Gynecology, University of Utah; Department of Cardiology, Cedars Sinai Medical Center; Department of Neurology, University of Pittsburgh; Department of Obstetrics & Gynecology, Columbia University; Department of Obstetrics & Gynecology, University of California, Irvine; Departments of Health, Sport, & Human Physiology and Epidemiology, University of Iowa; Departments of Obstetrics, Gynecology & Reproductive Sciences, Epidemiology, and Clinical and Translational Sciences, University of Pittsburgh

**Keywords:** Life’s Essential 8, adverse pregnancy outcomes, maternal cardiovascular health, nuMoM2b-HHS

## Abstract

**Background:** Individual health behaviors are associated with pregnancy outcomes, but their joint effects are rarely considered and their associations with health beyond pregnancy are uncertain. We aimed to examine associations between combinations of first trimester health behaviors with adverse pregnancy outcomes (APOs) and blood pressure 2-7 years after delivery, and to estimate the proportion of associations with later blood pressure that were mediated by APOs.

**Methods:** Participants in the nuMoM2b and follow-up Heart Health Study prospective cohort were included. Physical activity, diet, sleep duration, and smoking were scored using the Life’s Essential 8 framework. APOs were prospectively abstracted from medical records and included hypertensive disorders of pregnancy, preterm birth, gestational diabetes, small-for-gestational-age birth, or stillbirth. Latent profiles of health behaviors were constructed with structural equation modelling. Models were built using first trimester cardiovascular health scores on a scale of 0-100 for each of the four health behaviors. Risk of APOs and incident hypertension, and differences in continuous blood pressure 2-7 years after delivery based on behavioral profiles were assessed with Poisson or linear regression. Mediation analysis examined the proportion of associations between behavioral profiles and blood pressure mediated by APOs.

**Results:** Among 8,700 nulliparas, four behavioral profiles were identified: Healthiest Behaviors (36%), Healthy Activity/Sleep with Poor Diet/Smoking (21%), Healthy Sleep Only (32%), and Least Healthy Behaviors (11%). Forty-nine percent (4,497) returned for blood pressure assessment 2-7 years after delivery. Adjusted risk of APOs was 12-23% higher across the less healthy behavioral profiles compared to the Healthiest Behaviors profile. Further, adjusted systolic and diastolic blood pressures were 0.86 and 1.04 mmHg higher, respectively, 2-7 years after delivery in those with less healthy behavioral patterns compared to the Healthiest Behaviors profile. Eleven percent of the association between behavioral profiles and diastolic blood pressure was mediated by APOs. Rates of incident hypertension were not significantly different across behavioral profiles.

**Conclusions:** Less healthy combined health behavior profiles in early pregnancy were associated with APOs and higher blood pressure 2-7 years after delivery. The observed associations with later blood pressure were predominately direct effects, with limited mediation through APOs.

## Introduction

Pregnancy is considered a cardiovascular stress test that can provide an early warning for future risk of cardiovascular disease.^1^ Adverse pregnancy outcomes (APOs), including hypertensive disorders of pregnancy, preterm birth, or gestational diabetes are associated with a more than two-fold higher risk of developing hypertension within seven years of delivery^2–7^, suggesting that shared antecedents may exist. However, accurately identifying those who will develop APOs remains elusive, and prevention strategies are limited.

The American Heart Association’s Life’s Essential 8 framework for cardiovascular health (CVH)^8^ includes health factors (blood glucose, lipids, body mass index [BMI], and blood pressure) and health behaviors (physical activity, diet quality, sleep duration, and nonsmoking) that can be assessed in clinical settings and provides a metric to help identify those at higher risk of APOs and later cardiovascular disease.^1^ An early-pregnancy atherogenic profile derived from the health factor components has been associated with both APOs and incident hypertension 2-7 years after delivery in nulliparous individuals.^9^ Separate modifiable health behavior components also impact pregnancy health^10–16^, but importantly, health behaviors do not exist in isolation and adherence to multiple healthy behaviors can have additive beneficial effects.^17–19^ Co-occurring profiles of behavioral patterns during pregnancy have yet to be evaluated as precursors of APOs, and it is unknown whether combined health behaviors in early pregnancy are related to long-term cardiovascular outcomes such as hypertension.

Furthermore, there is a growing body of evidence demonstrating that, while APOs are associated both with poor early-pregnancy CVH and later cardiovascular disease, early pregnancy health factors themselves are related to future CVD risk even among those without APOs.^5,9,20^ While this has been demonstrated for clinical health factors such as BMI, blood pressure, lipids, and glucose, the contributions of health behaviors to these associations are uncertain. The Nulliparous Pregnancy Outcomes: Monitoring Mothers-to-be prospective cohort and its follow-up Heart Health Study (nuMoM2b-HHS) includes early-pregnancy assessment of health behaviors, prospective collection of rigorously defined APO diagnoses, and postpartum follow-up of cardiovascular risk metrics. Therefore, the objectives of this secondary analysis were: 1) to identify profiles of health behaviors in early pregnancy, 2) examine the associations of these profiles with incident APOs and maternal blood pressure several years after delivery, and 3) determine whether APOs mediated associations between behavioral profiles and later maternal blood pressure.

## Methods

### Study Design and Population

Study protocols for the nuMoM2b-HHS prospective cohort have been described previously.^21,22^ Briefly, 10,038 nulliparous pregnant individuals were recruited between 2010-2013. Participants were enrolled in the first trimester and followed through delivery at eight clinical sites across the United States. Participants were eligible if they were 6 weeks+ 0 days to 13 weeks+ 6 days pregnant with a singleton gestation and had no prior pregnancy that lasted past 20 weeks. In-person postpartum follow-up occurred 2-7 years following the index pregnancy (n=4,508). Ethical approval was obtained at each clinical site and participants provided written informed consent. Participants included in this analysis delivered at or after 20 weeks+ 0 days of gestation; those included in the APO analysis had complete pregnancy outcomes data, while those included in the analysis of future blood pressure returned for an in-person follow-up assessment and had complete blood pressure data. See **Figure 1** for the study flow diagram.

**Figure 1.**
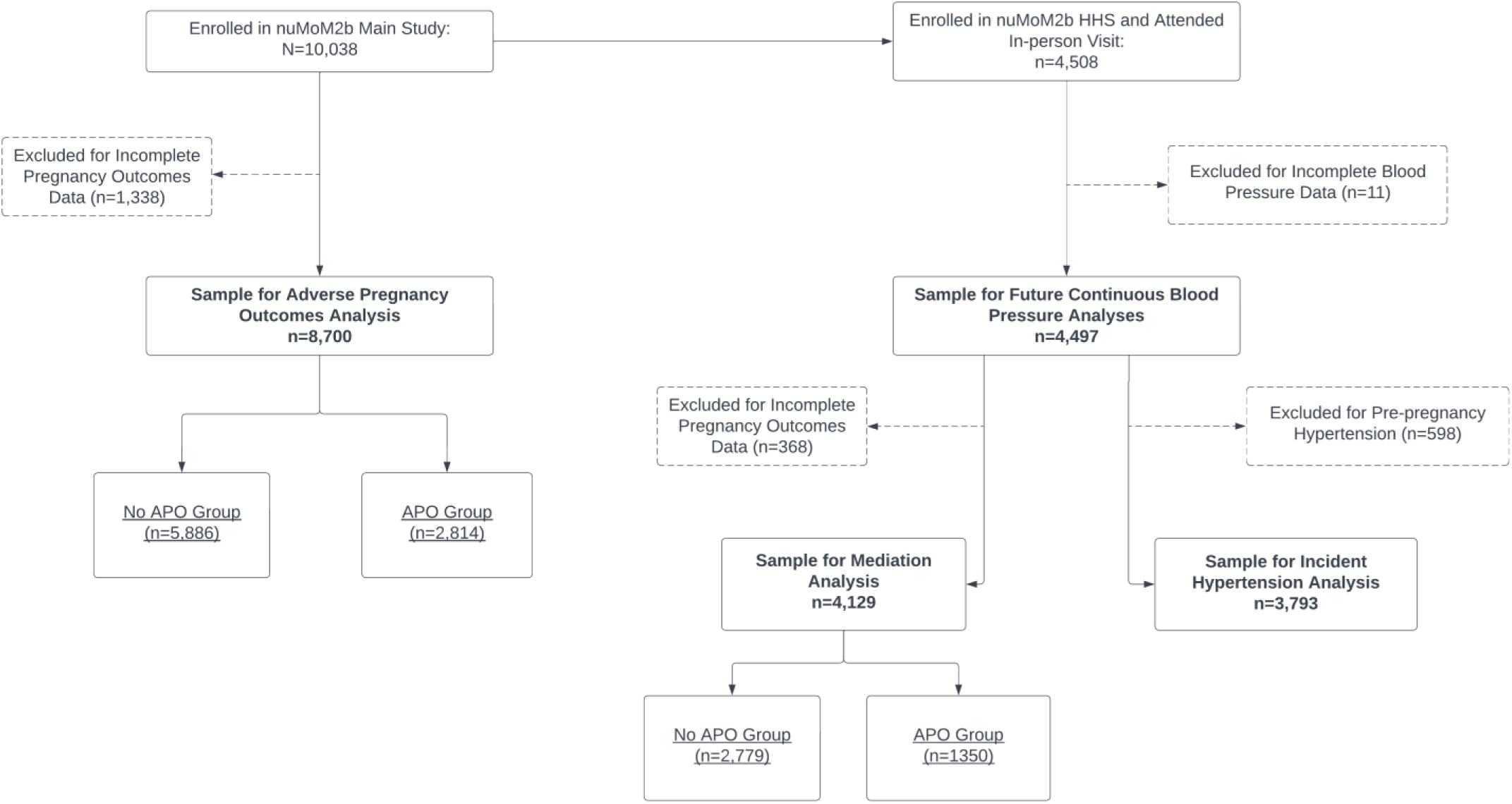
Study flow diagram. APO: adverse pregnancy outcome; nuMoM2b-HHS1: Nulliparous Pregnancy Outcomes: Monitoring Mothers-to-be Heart Health Study.

### Health Behaviors

Health behaviors were self-reported at the first trimester assessment. Leisure-time physical activity type, duration, and frequency were assessed using a questionnaire adapted from the Behavior Risk Factor Surveillance System^23,24^, converted to metabolic equivalents of the task using the Compendium of Physical Activities, 2^nd^ edition^25^, and quantified as minutes per week of moderate-to-vigorous physical activity (MVPA). Smoking history and exposure to secondhand smoke in the home were self-reported by questionnaire. Nightly hours of sleep duration were self-reported separately for weekdays and weekends; the mean was computed to calculate average nightly sleep duration over a 1-week period. Diet quality was assessed using a modified Block Food Frequency Questionnaire^26,27^ and converted to a Healthy Eating Index (HEI) score. Physical activity, smoking, and sleep questions referenced the first trimester, while the diet questionnaire referenced the three months immediately preceding conception.

Each behavioral component was assigned a score from 0-100 using the American Heart Association’s Life’s Essential 8 algorithm (**Table S1**). Scores ≥80 indicate “ideal” health, while scores of 50-79 indicate “moderate” health and scores <50 indicate “poor” health.^28^ Component scores for physical activity ranged from 0 (no weekly MVPA) to 100 (≥150 minutes/week MVPA) with intermediate scores for values between 0-150 minutes/week. HEI score percentiles were used to calculate the diet score. For example, 0 points corresponded to HEI <25^th^ percentile, while 100 points corresponded to HEI ≥95^th^ percentile. Sleep scores accounted for the U-shaped association between sleep duration and cardiovascular health, in which both low and high sleep durations are adverse.^29^ A sleep score of 100 corresponded to 7-9 hours of sleep, with lower scores for fewer or greater hours of nightly sleep. Finally, the smoking score was adapted for pregnancy from the standard scoring algorithm. A score of 0 indicated current smoking during pregnancy, while a score of 100 indicated never smoking tobacco; intermediate scores were assigned for former smokers. In accordance with the standard scoring algorithm, twenty points were subtracted in cases of exposure to secondhand smoke in the home for all except current smokers.^8^

### Adverse Pregnancy Outcomes

Pregnancy outcomes were determined through medical record abstraction. We used a binary composite APO variable, defined as presence of any of the following: hypertensive disorders of pregnancy (>140 mmHg systolic or >90 mmHg diastolic at or after 20 gestational weeks with or without proteinuria, end-organ involvement, and/or HELLP syndrome), gestational diabetes, preterm birth (<37 weeks of gestation at delivery), small-for-gestational-age birth (<5^th^ percentile using Alexander curves), or stillbirth.

### Blood Pressure 2-7 Years After Delivery

Resting blood pressure was measured at the in-person nuMoM2b-HHS1 visit, occurring 2-7 years following the index pregnancy, using a standardized protocol.^22^ Following a seated rest, blood pressure was measured three times using an automated device (Omron Healthcare Incorporated, Lake Forest, IL); the last two measurements were averaged and used for analysis. Incident hypertension was defined as new onset of stage 1 hypertension (≥130 mmHg systolic or ≥80 mmHg diastolic) or the use of blood pressure lowering medication at follow-up. Those with measured first trimester stage 1 hypertension were excluded from incident hypertension analyses. Finally, we examined associations with continuous values of systolic and diastolic blood pressure at follow-up.

### Covariates and Demographic Characteristics

Questionnaires and interviews assessed participant age at study enrollment, income, education, insurance, perceived racial discrimination (Krieger Discrimination Scale^30^, categorized as none, moderate, or high), and the social construct of self-reported racial and ethnic identity. For those self-reporting anti-hypertensive medication usage at the follow-up visit (n=117), we added 10/5 mmHg to measured systolic/diastolic blood pressures, respectively.^31^ Height and weight were measured at the first trimester study visit using standardized protocols and were converted into body mass index (BMI) in kg/m^2^. Self-reported pre-pregnancy diabetes mellitus was assessed at the first trimester visit by interview. Resting blood pressure was measured at the first trimester study visit using the same protocol described above.

### Statistical Analyses

Latent (unobserved) classes of combinations of the four health behaviors were constructed empirically using the “gsem” command in Stata, which can compute the models using continuous values. Models were built using first trimester CVH scores on a scale of 0-100 for each of the four health behaviors (physical activity, diet, sleep duration, and smoking) based on the full nuMoM2b sample (N=10,038), and the best-fitting model was selected based on values of Akaike/Bayesian Information Criterion, sample size in each class, and model interpretability; posterior probabilities were also assessed.

We assessed the risk of having an APO among behavioral latent profiles using Poisson regression with robust error variance. Crude models were first adjusted for participant characteristics including age, insurance status, income, education, and perceived racial discrimination; a final model also included first trimester blood pressure and BMI, which are health factors that may both mediate and confound the association between health behaviors and our outcomes.^32,33^

Next, we estimated the risk of incident stage 1 hypertension 2-7 years following delivery associated with early pregnancy behavioral profiles using Poisson regression with robust error variance. In addition, we tested the association between behavioral profiles and continuous blood pressure 2-7 years after delivery; separate models were tested for systolic and diastolic blood pressure using linear regression. Individuals with early-pregnancy stage 1 hypertension (n=598, 13%) were excluded from the incident hypertension models but were included in analyses of APOs and continuous blood pressure values. Crude associations for both incident hypertension and continuous blood pressure values were first adjusted for the time elapsed between delivery and the follow-up assessment and first-trimester participant sociodemographic characteristics. A final model added covariate adjustment for first trimester blood pressure and BMI.

Next, we conducted a mediation analysis to estimate the proportion of any significant associations between early-pregnancy health behaviors and blood pressure outcomes 2-7 years after delivery that were mediated by APOs. We used a multiplicative model with an interaction term between behavioral profile and APO status (modeled as a count outcome using Poisson regression). Decomposition of the total effects included estimations of how much early pregnancy health behaviors directly affected later blood pressure (natural direct effect) and how much of the effect was explained by APOs (natural indirect effect); we also estimated the proportions of the total effect mediated by APOs.

Crude models were adjusted using the framework depicted in **Figure S1**.

Finally, we conducted the following sensitivity analyses: first, we evaluated associations between early-pregnancy health behaviors and the APO and continuous blood pressure outcomes when excluding those with early-pregnancy stage 1 hypertension or diabetes. Second, the clinical cutoff for hypertension in pregnancy is ≥140 mmHg systolic and/or ≥90 mmHg diastolic and the pregnancy measurements were taken prior to the implementation of revised blood pressure categorizations^34^, so we examined whether associations with incident hypertension were different using incidence of stage 2 hypertension. All analyses were conducted using Stata version 18 (StataCorp, College Station, TX) and significance was accepted at p<0.05.

## Results

In total, 8,700 participants (mean ± SD 26.9 ± 5.6 years, 26.2 ± 6.3 kg/m^2^ BMI, 40% underrepresented race or ethnicity) with complete pregnancy outcomes data were included in the APO analysis (**Table 1**). Model fit criteria for latent classes of early-pregnancy health behaviors found a four-class solution to be the most appropriate (AIC = 343338.1, BIC = 343504.0, Entropy = 0.85; **Table S2**). Participants were categorized as engaging in Healthiest Behaviors (n=3,158, 36.3%), Healthy Activity/Sleep with Poor Diet/Smoking (n=1,807, 20.8%), Healthy Sleep Only (n=2,804, 32.2%), or Least Healthy Behaviors (n=931, 10.7%) (**Figure 2**). Participants in the Healthiest Behaviors profile were more likely to be older, have a lower BMI, have higher education and income, self-identify as non-Hispanic White, have lower systolic and diastolic blood pressure in the first trimester of pregnancy, and have a lower prevalence of stage 1 hypertension at baseline compared to those in the other behavioral profiles.

**Figure 2.**
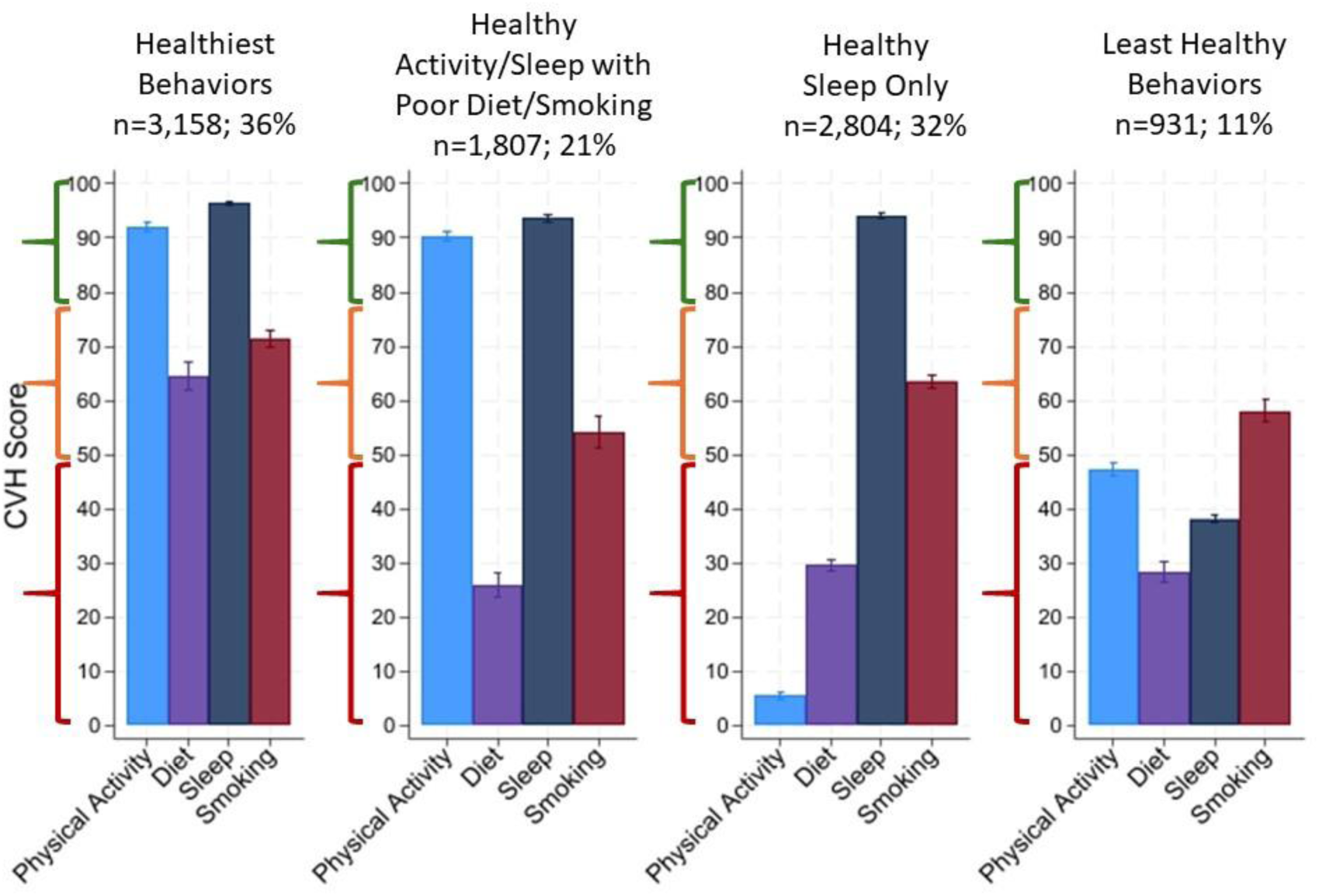
Health behavior cardiovascular health component scores by behavioral profiles. Mean (SD) cardiovascular health (CVH) scores for each health behavior component within each behavioral profile. Higher score indicates more ideal CVH; scores <50 are “poor” CVH; 50-79 are “moderate” CVH, and scores ≥80 are “high” CVH as indicated by markers on the y-axis.

**Table 1.** Participant characteristics by health behavior latent profiles.

|  | Healthiest Behaviors<br>(n=3,158;<br>36%) | Healthy Activity/Sleep<br>with Poor Diet/Smoking<br>(n=1,807;<br>21%) | Healthy Sleep Only<br>(n=2,804;<br>32%) | Least Healthy Behaviors<br>(n=931;<br>11%) | *P* |
| --- | --- | --- | --- | --- | --- |
| **Maternal age (years)** | 29.0 ± 4.9 | 26.2 ± 5.7 | 25.8 ± 5.6 | 24.4 ± 5.5 | <0.001 |
| **Body mass index (kg/m<sup>2</sup>)** | 25.1 ± 5.1 | 27.1 ± 6.9 | 26.8 ± 6.5 | 27.1 ± 7.3 | <0.001 |
| **Maternal Race** |  |  |  |  | <0.001 |
| American Indian | 2 (<1%) | 2 (<1%) | 0 (0%) | 3 (<1%) |  |
| Asian | 167 (5%) | 51 (3%) | 107 (4%) | 28 (3%) |  |
| Black non-Hispanic | 183 (6%) | 298 (16%) | 481 (17%) | 203 (22%) |  |
| Hispanic | 338 (11%) | 275 (15%) | 659 (24%) | 226 (24%) |  |
| Multiracial | 103 (3%) | 87 (5%) | 111 (4%) | 48 (5%) |  |
| Native Hawaiian | 9 (<1%) | 11 (<1%) | 8 (<1%) | 5 (<1%) |  |
| White non-Hispanic | 2,334 (74%) | 1,074 (59%) | 1,422 (51%) | 415 (45%) |  |
| Not available | 22 (<1%) | 9 (<1%) | 16 (<1%) | 3 (<1%) |  |
| **Educational Attainment** |  |  |  |  | <0.001 |
| Less than high school | 58 (2%) | 186 (10%) | 314 (11%) | 142 (15%) |  |
| Completed high school or GED | 98 (3%) | 254 (14%) | 455 (16%) | 209 (22%) |  |
| Some college | 362 (11%) | 429 (24%) | 670 (24%) | 243 (26%) |  |
| Associate or technical degree | 261 (8%) | 198 (11%) | 324 (12%) | 99 (11%) |  |
| Completed college | 1156 (37%) | 448 (25%) | 641 (23%) | 146 (16%) |  |
| Degree work beyond college | 1219 (39%) | 292 (16%) | 394 (14%) | 92 (10%) |  |
| **Percentage of Federal Poverty Level** |  |  |  |  | <0.001 |
| <100% | 177 (6%) | 284 (20%) | 452 (22%) | 194 (31%) |  |
| 100-200% | 263 (9%) | 250 (18%) | 387 (18%) | 115 (18%) |  |
| >200% | 2459 (85%) | 887 (62%) | 1253 (60%) | 318 (51%) |  |
| **Insurance Payor** |  |  |  |  | <0.001 |
| Commercial | 2000 (64%) | 892 (50%) | 1371 (49%) | 334 (36%) |  |
| Medicare/Medicaid | 296 (9%) | 583 (32%) | 990 (36%) | 448 (48%) |  |
| Military/Other | 39 (1%) | 44 (2%) | 42 (2%) | 24 (3%) |  |
| Two or more sources | 730 (23%) | 219 (12%) | 296 (11%) | 91 (10%) |  |
| None (personal income) | 80 (3%) | 56 (3%) | 75 (3%) | 27 (3%) |  |
| **1<sup>st</sup> Trimester Systolic BP (mmHg)** | 108.6 ± 10.4 | 110.0 ± 11.0 | 109.2 ± 11.4 | 108.8 ± 11.5 | <0.001 |
| **1<sup>st</sup> Trimester Diastolic BP (mmHg)** | 67.0 ± 8.1 | 67.5 ± 8.5 | 67.0 ± 8.5 | 66.4 ± 8.7 | 0.014 |
| **1<sup>st</sup> Trimester Stage 1 Hypertension** | 332 (10.7%) | 257 (14.5%) | 369 (13.5%) | 122 (13.4%) | <0.001 |
| **1<sup>st</sup> Trimester Stage 2 Hypertension** | 35 (1.1%) | 36 (2.0%) | 47 (1.7%) | 18 (2.0%) | 0.058 |
| **Composite APO** | 882 (28%) | 635 (35%) | 948 (34%) | 349 (37%) | <0.001 |

*Reported as mean ± standard deviation or frequency and percentage, as appropriate.*
*Differences among behavioral latent classes tested using one-way analysis of variance for continuous variables or Chi-squared tests for categorical variables. APO: adverse pregnancy outcomes; BP: blood pressure; GED: general education diploma.*

Early pregnancy behavioral profiles were significantly associated with risk of APOs (**Table 2**), with those in less healthy profiles having a 21-34% higher unadjusted risk compared to those in the Healthiest Behaviors profile [overall Chi-squared (3) = 33.28, p<0.001]. These significant associations remained following adjustment for sociodemographic characteristics, ranging from a 12-23% higher risk. These associations persisted in the Healthy Activity/Sleep with Poor Diet/Smoking (15% higher, 95% confidence interval 2-30%) and Least Healthy Behaviors (18% higher, 95% confidence interval 1-38%) profiles upon further adjustment for early pregnancy blood pressure and BMI. A full summary of individual APO diagnoses by behavioral profiles is shown in **Table S3.**

**Table 2.** Incident rate ratios (IRR) of adverse pregnancy outcomes by behavioral profiles.

| Group | Model 1 IRR<br>(95% CI) | *P* | Model 2 IRR<br>(95% CI) | *P* | Model 3 IRR<br>(95% CI) | *P* |
| --- | --- | --- | --- | --- | --- | --- |
| Healthiest Behaviors | 1.00 (ref) | - | 1.00 (ref) | - | 1.00 (ref) | - |
| Healthy Activity/Sleep with Poor Diet/Smoking | **1.26 (1.16, 1.37)** | **<0.001** | **1.20 (1.09, 1.32)** | **<0.001** | **1.15 (1.04, 1.27)** | **0.004** |
| Healthy Sleep Only | **1.21 (1.12, 1.31)** | **<0.001** | **1.12 (1.02, 1.22)** | **0.016** | 1.09 (0.99, 1.19) | 0.068 |
| Least Healthy Behaviors | **1.34 (1.21, 1.48)** | **<0.001** | **1.23 (1.09, 1.39)** | **0.001** | **1.18 (1.04, 1.34)** | **0.010** |

*Model 1: unadjusted; Model 2: adjusted for maternal age, education, insurance, income, and perceived racial discrimination; Model 3: additionally adjusted for first trimester blood pressure and body mass index. Estimates modeled with Poisson regression with robust error variance. Bold font indicates significant difference ( $p < 0.05$ ).*

Of the original cohort, 4,497 returned for post-delivery follow-up at 3.2 ± 0.9 years (range, 2-7 years) following the index pregnancy and had complete blood pressure data. Given the substantial overlap in APO risk among the three less favorable behavioral profiles, we dichotomized the groups as Healthiest Behaviors and Less Healthy Behaviors (including the Healthy Activity/Sleep with Poor Diet/Smoking, Healthy Sleep Only, and Least Healthy Behaviors profiles) for analyses relating to maternal blood pressure 2-7 years following delivery. Incident stage 1 hypertension was found in 18.1% of the sample who participated in the follow-up visit and did not have early-pregnancy stage 1 hypertension (688 of 3,793 participants), with no significant differences in incidence between early-pregnancy behavioral profiles (Healthiest Behaviors profile: 247 of 1,429, 17.3%; other three Less Healthy Behaviors profiles combined: 441 of 1,923, 18.7%, IRR = 1.08 [95% confidence interval 0.94 – 1.24, p=0.290], **Table 3**).

**Table 3.** Association of incident hypertension and continuous blood pressure by behavioral profiles.

| Group | Model 1 | P | Model 2 | P | Model 3 | P |
| --- | --- | --- | --- | --- | --- | --- |
| **Incident Hypertension [IRR (95% CI)]** |  |  |  |  |  |  |
| Healthiest Behaviors | 1.00 (ref) | - | 1.00 (ref) | - | 1.00 (ref) | - |
| Less Healthy Behaviors | 1.08 (0.94, 1.24) | 0.290 | 1.01 (0.85, 1.19) | 0.906 | 0.94 (0.80, 1.10) | 0.442 |
| **Systolic Blood Pressure, mmHg [β coefficient (95% CI)]** |  |  |  |  |  |  |
| Healthiest Behaviors | Reference | - | Reference | - | Reference | - |
| Less Healthy Behaviors | **1.42 (0.72, 2.12)** | **<0.001** | **0.86 (0.06, 1.66)** | **0.035** | 0.54 (-0.23, 1.32) | 0.170 |
| **Diastolic Blood Pressure, mmHg [β coefficient (95% CI)]** |  |  |  |  |  |  |
| Healthiest Behaviors | Reference | - | Reference | - | Reference | - |
| Less Healthy Behaviors | **1.49 (0.87, 2.10)** | **<0.001** | **1.04 (0.33, 1.74)** | **0.004** | 0.54 (-0.14, 1.22) | 0.118 |

*Model 1: unadjusted; Model 2: adjusted for time elapsed between visits, maternal age, education, insurance, income, and perceived racial discrimination; Model 3: additionally adjusted for first trimester blood pressure and body mass index. Incident hypertension modeled with Poisson regression with robust error variance; continuous blood pressure estimates modeled with linear regression. Bold font indicates significant difference ( $p < 0.05$ ). IRR: incidence rate ratio.*

Crude linear regression results for continuous blood pressure found differences that were retained after adjustment for visit date and sociodemographic factors (**Table 3**), wherein participants in the combined less favorable behavior profiles had 0.86 mmHg higher systolic blood pressure (p=0.035) and 1.04 mmHg higher diastolic blood pressure (p=0.004) compared to the Healthiest Behaviors profile. After additional adjustment for first trimester blood pressure and BMI, these associations were no longer significant.

In analyses examining the mediation by APOs of the association between early pregnancy behavioral profiles and long-term blood pressure, significant total, natural direct, and natural indirect effects were found in crude models for both diastolic and systolic blood pressure (**Table 4**) which were retained upon adjustment for visit date and sociodemographic factors. When the models were additionally adjusted for early pregnancy blood pressure and BMI, total and natural direct effects remained significant for diastolic blood pressure while associations with systolic blood pressure were no longer significant. The proportion of the association mediated through APOs was 11% for diastolic blood pressure in Model 2 (p=0.034) and was nonsignificant for systolic blood pressure.

**Table 4.** Effect decomposition of the association between behavioral profiles and future blood pressure mediated by adverse pregnancy outcomes.

|  | Effect Decomposition |  |  |  |
| --- | --- | --- | --- | --- |
|  | Natural Indirect<br>Effect, *P* | Natural Direct<br>Effect, *P* | Total Effect,<br>*P* | Proportion<br>mediated, *P* |
| **Systolic Blood Pressure (mmHg)** |  |  |  |  |
| Model 1 | **0.23, p<0.001** | **1.40, p<0.001** | **1.62, p<0.001** | **14%, p=0.002** |
| Model 2 | **0.16, p=0.023** | **0.97, p=0.018** | **1.13, p=0.007** | 14%, p=0.054 |
| Model 3 | 0.09, p=0.110 | 0.69, p=0.084 | 0.78, p=0.055 | 11%, p=0.183 |
| **Diastolic Blood Pressure (mmHg)** |  |  |  |  |
| Model 1 | **0.21, p<0.001** | **1.52, p<0.001** | **1.74, p<0.001** | **12%, p=0.001** |
| Model 2 | **0.15, p=0.022** | **1.24, p=0.001** | **1.38, p<0.001** | **11%, p=0.034** |
| Model 3 | 0.07, p=0.113 | **0.80, p=0.024** | **0.87, p=0.015** | 8%, p=0.154 |

*Model 1: unadjusted; Model 2: adjusted for maternal age, education, insurance, income, and perceived racial discrimination; Model 3: additionally adjusted for first trimester blood pressure and body mass index. Bold font indicates significant difference (p<0.05).*

Sensitivity analyses excluding participants with pre-pregnancy stage 1 hypertension and/or diabetes from the models estimating APO risk and associations with continuous blood pressure 2-7 years after delivery did not change the direction or interpretation of our results (**Tables S4 and S5**). Similarly, rates of incident stage 2 hypertension were not different between those with healthy versus less healthy behaviors (**Table S6**).

## Discussion

In this diverse prospective cohort of individuals enrolled during a first pregnancy we found four distinct first-trimester health behavior profiles that were associated with risk of subsequent APOs. Those with less healthy behavioral profiles were 12-23% more likely to experience an APO than those with the healthiest behavioral profile. Further, less healthy early-pregnancy behavioral profiles were associated with higher blood pressure 2-7 years following delivery. Mediation analysis suggested that these effects were predominantly direct, rather than mediated through APOs. Our findings indicate that healthy behavior combinations in early pregnancy exhibit statistically significant associations with APOs and that behaviors in early pregnancy may be important for long-term cardiovascular risk whether or not an APO occurs.

By utilizing the Life’s Essential 8 framework, we were able to construct latent profiles of healthy behaviors (physical activity, diet quality, sleep duration, and nonsmoking) in early pregnancy. This allowed us to leverage naturally occurring covariance in healthy behaviors during pregnancy, rather than forcing disaggregation. We found that lower adherence to healthy behaviors was associated with greater risk of APOs. This was particularly evident in the Least Healthy Behaviors profile (comprised of individuals with “poor” adherence to physical activity, diet quality, and sleep duration behaviors and “moderate” adherence to nonsmoking behavior), in which the adjusted risk ratio was highest. These results suggest that healthy behaviors in early pregnancy may be additive, and that “poor” adherence to at least two healthy behaviors in early pregnancy is related to elevated risk of APOs. Our findings are in contrast to those from a small cohort (n=202)^15^ in which individual health behaviors, but not a composite score, were associated with features of placental health. This discrepancy may be attributable to cohort differences, distinct scoring algorithms, or study outcomes.

In addition, associations between behavioral profiles and APOs were independent of first trimester blood pressure and BMI. These findings suggest that engaging in healthy behaviors is an important contributor to pregnancy outcomes for all pregnant individuals, regardless of these clinical health factors in early pregnancy. Our results support existing evidence that healthy behaviors are associated with more favorable pregnancy outcomes^10–16,35^, and add important knowledge about the combined effects of multiple healthy behaviors. Furthermore, these results emphasize the importance of healthy behaviors even for those with favorable early-pregnancy blood pressure and BMI.

It is also important to note sociodemographic differences among participants in each behavioral profile in our study. Those in profiles with the lowest adherence to healthy behaviors and the highest risk of APOs were likely impacted by structural factors that may have influenced the ability to adhere to and the interplay between the healthy behaviors (e.g., neighborhood factors that may influence access to both physical activity and healthy diet options). While we adjusted for sociodemographic factors, future research should investigate the drivers of these behavioral profiles and identify areas for intervention, such as enhancement of the built environment.

We found no significant association between behavioral profiles and incident hypertension 2-7 years after delivery. However, when examining associations among behavioral profiles and continuous blood pressure over this time, we found that those with less healthy behavioral profiles had 0.86 mmHg higher systolic blood pressure and 1.04 mmHg higher diastolic blood pressure 2-7 years following delivery than those in the Healthiest Behaviors profile. Our mediation analysis suggests that health behaviors may play a direct role in longer-term continuous blood pressure levels in the years following delivery, with a small but significant 11% of the effect on diastolic blood pressure mediated through APOs. This is similar to two prior studies in the nuMoM2b-HHS cohort. In the first, the association of an early-pregnancy atherogenic profile of poor lipids, C-reactive protein, and insulin with postpartum incident hypertension was partially mediated by APOs.^9^ In the second, hypertensive disorders of pregnancy mediated a small but statistically significant proportion of the association between early pregnancy obesity and incident hypertension.^20^ Taken together, the results of our study and this prior work suggest that early pregnancy CVH components are robustly associated with future maternal cardiovascular risk, with a small proportion of these associations mediated by APO occurrence (7-14% across these studies). Our findings point to APOs as possible contributors to higher future blood pressure, while behavioral profiles in early pregnancy are important whether or not an APO occurs.

While 0.86-1.04 mmHg higher blood pressure over a mean follow-up of three years is not a clinically large difference, small increases in blood pressure in the reproductive years can compound cardiovascular risk over time.^36,37^ We found that adjusting for first trimester blood pressure and BMI attenuated our results, suggesting that these clinical factors may themselves mediate the association between health behaviors in early pregnancy and later blood pressure. There are multiple possible explanations of this finding. First, it is possible that early pregnancy blood pressure confounded the association with blood pressure in the years following delivery (i.e., those with higher blood pressure following delivery simply started with higher blood pressure in early pregnancy). Second, it is likely that blood pressure and BMI were in the causal pathway between early pregnancy health behaviors and blood pressure 2-7 years after delivery (i.e., those with healthier behaviors had more favorable early-pregnancy blood pressure and BMI due to the engagement in those healthy behaviors).^32,33^ Given this potential mediating effect, we chose to report Model 2 as our primary results, which did not include early-pregnancy blood pressure and BMI. In either case, these findings emphasize the potential importance of maximizing pre-conception and early pregnancy health in order to limit the development of CVD risk factors in this critical peripartum window^1^, and suggest that engaging in healthy behaviors may be a strategy to achieve this. Future interventional research is needed to determine whether improving engagement in healthy behaviors in the prenatal period is an effective means to improve long-term maternal health.

Pregnancy is a time of near-universal healthcare access, and motivation may be high to engage in behavior change.^38,39^ Our results suggest that modifiable health behaviors group together in distinct ways, and that risks of multiple unhealthy behaviors compound one another and may be related to health beyond pregnancy. Notably, we did not see large differences in risk between our three less healthy behavioral profile groups, but each group with two or more behaviors classified as “poor” were at higher risk. This suggests that behavior change interventions may employ strategies to improve adherence to whichever health behavior is the most easily modifiable for the individual rather than targeting the adoption of a particular behavioral profile. Early pregnancy may be an ideal time to assess behavioral profiles and engage individuals in behavior change.

## Strengths, Limitations, and Future Directions

This study has several strengths. We utilized a longitudinal approach to the reproductive years by examining early-pregnancy behavioral profile exposures and followed participants through pregnancy and several postpartum years. Our large, diverse prospective cohort of people experiencing a first birth captured well-documented APOs. In addition, we were able to identify four latent health behavior profiles, demonstrating for the first time how different behavioral combinations in early pregnancy are related to perinatal and long-term maternal health. Those who returned for a postpartum visit 2-7 years after delivery were demographically similar to the full pregnancy cohort^2^, suggesting these results may be generalizable to our sample at large.

A limitation of this study included the self-report of health behaviors. However, given that these behaviors were reported prior to the outcomes occurring, recall bias or other measurement error would likely be random and therefore biased toward the null. In addition, the diet questionnaire referenced the 3 months prior to conception, which may not have accurately captured first trimester dietary patterns. Despite these limitations, our strong posterior probabilities of latent class membership suggest that the behavioral profiles are distinct and any bias is likely nondifferential. Furthermore, our activity questionnaire was limited to leisure-time physical activities. Future investigations should examine whether behavioral patterns are different among those with differing levels of occupational activity. Finally, health behaviors may have shifted between delivery and the postpartum follow-up assessment; future research should examine whether behavioral profiles are consistent after childbirth and whether more recent behavioral profiles explain additional variability in cardiovascular risk measures.

## Conclusions

Our results suggest that co-occurring combinations of modifiable health behaviors are important and are related to pregnancy outcomes and blood pressure years following delivery. These findings add to the growing body of literature indicating that health in early pregnancy is a strong contributor to both pregnancy health and CVH following pregnancy. Importantly, health behaviors also had direct associations with blood pressure in the years following delivery and thus are important for those with and without pregnancy complications. Improving health behaviors in the prenatal context shows promise for reducing maternal risk during and possibly beyond pregnancy.

## Data Availability

Publicly available data related to this study are available on the DASH data and specimen hub.

## Non-standard Abbreviations and Acronyms

APO: adverse pregnancy outcome
BMI: body mass index
CVH: cardiovascular health
HEI: Healthy Eating Index
HELLP: hemolysis, elevated liver enzymes, and low platelets
MVPA: moderate-to-vigorous physical activity
nuMoM2b-HHS: Nulliparous Pregnancy Outcomes: Monitoring Mothers-to-be Heart Health Study

## Acknowledgements

Grant funding from the Eunice Kennedy Shriver National Institute of Child Health and Human Development (NICHD): U10 HD063036; U10 HD063072; U10 HD063047; U10 HD063037; U10 HD063041; U10 HD063020; U10 HD063046; U10 HD063048; and U10 HD063053. In addition, support was provided by Clinical and Translational Science Institutes: UL1TR001108 and UL1TR000153. Cooperative agreement funding from the National Heart, Lung, and Blood Institute and the Eunice Kennedy Shriver National Institute of Child Health and Human Development: U10-HL119991; U10-HL119989; U10-HL120034; U10-HL119990; U10-HL120006; U10-HL119992; U10-HL120019; U10-HL119993; U10-HL120018, and U01HL145358; and the National Center for Advancing Translational Sciences through UL-1-TR000124, UL-1-TR000153, UL-1-TR000439, and UL-1-TR001108; and the Barbra Streisand Women’s Cardiovascular Research and Education Program, and the Erika J. Glazer Women’s Heart Research Initiative, Cedars-Sinai Medical Center, Los Angeles. The Sedentary Behavior and Cardiovascular Health in Young Women ancillary study is supported by NHLBI R01HL158652. ACK is supported under NHLBI T32HL083825.

## Supplemental Tables

**Table S1.** Cardiovascular Health Scoring algorithm for health behaviors in pregnancy. MVPA: moderate-to-vigorous physical activity; HEI: Healthy Eating Index.

|  | Value | Cardiovascular Health Score |
| --- | --- | --- |
| Physical Activity<br>(minutes<br>MVPA/week) | 0 | 0 |
|  | 1-29 | 20 |
|  | 30-59 | 40 |
|  | 60-89 | 60 |
|  | 90-119 | 80 |
|  | 120-149 | 90 |
|  | 150+ | 100 |
| Diet (HEI<br>percentile) | <25<sup>th</sup> | 0 |
|  | 25<sup>th</sup>-49<sup>th</sup> | 25 |
|  | 50<sup>th</sup>-74<sup>th</sup> | 50 |
|  | 75<sup>th</sup>-94<sup>th</sup> | 80 |
|  | ≥95<sup>th</sup> | 100 |
| Sleep Duration<br>(mean hours/night) | <4 | 0 |
|  | 4 - <5 | 20 |
|  | 5 - <6 or ≥10 | 40 |
|  | 6 - <7 | 70 |
|  | 9 - <10 | 90 |
|  | 7 - <9 | 100 |
| Smoking | Current smoking | 0 |
|  | Smoked within 3<br>months of conception | 25 |
|  | Former smoker, quit<br>prior to 3 months before<br>conception | 50 |
|  | Never smoker | 100 |
|  | Lives with current<br>smoker | Subtract 20 points<br>unless score already =<br>0 |

**Table S2.** Latent Class metrics.

| **Solution comparisons** | **AIC** | **BIC** |
| --- | --- | --- |
| 1 Class Solution | 355988.6 | 356046.3 |
| 2 Class Solution | 353844.9 | 353938.7 |
| 3 Class Solution | 345209.8 | 345339.6 |
| 4 Class Solution | 343338.1 | 343504.0 |
| 5 Class Solution | 343348.1 | 343550.1 |
| **4 Class Solution Characteristics** | **N (%)** | **Posterior probability** |
| Healthiest behaviors | 3,158<br>(36.5%) | 0.78 |
| Healthy activity/sleep<br>with poor diet/smoking | 1,807<br>(20.6%) | 0.73 |
| Healthy sleep only | 2,804<br>(32.3%) | 0.95 |
| Least healthy behaviors | 931 (10.7%) | 0.95 |
| Overall | 8,700 (100%) | 0.85 |

*AIC: Akaike's information criterion; BIC: Bayesian information criterion.*

**Table S3.** Individual adverse pregnancy outcome diagnoses by behavioral profiles.

|  | Healthiest Behaviors<br>(n=3,158) | Healthy Activity/Sleep with Poor Diet/Smoking<br>(n=1,807) | Healthy Sleep Only<br>(n=2,804) | Least Healthy Behaviors<br>(n=931) |
| --- | --- | --- | --- | --- |
| Composite APO<br>(n, %) | 882 (30%) | 635 (35%) | 948 (34%) | 349 (37%) |
| *Preterm Birth* | 237 (8%) | 192 (11%) | 292 (10%) | 108 (12%) |
| *Early Preterm Birth* | 65 (2%) | 57 (3%) | 109 (4%) | 38 (4%) |
| *Hypertensive Disorders of Pregnancy* | 389 (12%) | 290 (16%) | 406 (14%) | 159 (17%) |
| *Gestational Diabetes* | 117 (4%) | 84 (5%) | 148 (5%) | 47 (5%) |
| *Small for Gestational Age* | 327 (10%) | 216 (12%) | 353 (13%) | 122 (13%) |
| *Stillbirth* | 12 (0.4%) | 16 (0.9%) | 20 (0.7%) | 6 (0.6%) |

*APO: adverse pregnancy outcome. Early preterm birth defined as prior to 34 weeks of gestation at birth.*

**Table S4.** Incident rate ratios (IRR) of adverse pregnancy outcomes by behavioral profiles, excluding those with pre-pregnancy hypertension or diabetes.

| Group | Model 1 IRR<br>(95% CI) | *P* | Model 2 IRR<br>(95% CI) | *P* | Model 3 IRR<br>(95% CI) | *P* |
| --- | --- | --- | --- | --- | --- | --- |
| Healthiest Behaviors | 1.00 (ref) | - | 1.00 (ref) | - | 1.00 (ref) | - |
| Healthy Activity/Sleep with Poor Diet/Smoking | **1.23 (1.12, 1.36)** | **<0.001** | **1.18 (1.06, 1.32)** | **0.003** | **1.15 (1.03, 1.28)** | **0.015** |
| Healthy Sleep Only | **1.18 (1.08, 1.29)** | **<0.001** | 1.09 (0.98, 1.21) | 0.108 | 1.07 (0.96, 1.19) | 0.209 |
| Least Healthy Behaviors | **1.33 (1.19, 1.49)** | **<0.001** | **1.22 (1.06, 1.41)** | **0.006** | **1.21 (1.05, 1.39)** | **0.010** |

*Model 1: unadjusted; Model 2: adjusted for maternal age, education, insurance, income, and perceived racial discrimination; Model 3: additionally adjusted for first trimester blood pressure and body mass index. Estimates modeled with Poisson regression with robust error variance. Bold font indicates significant difference ( $p < 0.05$ ).*

**Table S5.** Association of incident hypertension and continuous blood pressure by behavioral profiles, excluding those with pre-pregnancy diabetes (incident hypertension) or pre-pregnancy diabetes or hypertension (continuous blood pressure).

| Group | Model 1 | *P* | Model 2 | *P* | Model 3 | *P* |
| --- | --- | --- | --- | --- | --- | --- |
| **Incident Stage Hypertension [IRR (95% CI)]** |  |  |  |  |  |  |
| Healthiest Behaviors | 1.00 (ref) | - | 1.00 (ref) | - | 1.00 (ref) | - |
| Less Healthy Behaviors | 1.07 (0.93, 1.23) | 0.363 | 1.01 (0.85, 1.19) | 0.938 | 0.93 (0.79, 1.10) | 0.412 |
| **Systolic Blood Pressure, mmHg [β coefficient (95% CI)]** |  |  |  |  |  |  |
| Healthiest Behaviors | Reference | - | Reference | - | Reference | - |
| Less Healthy Behaviors | **1.20 (0.50, 1.89)** | **0.001** | **0.83 (0.02, 1.65)** | **0.045** | 0.56 (-0.24, 1.36) | 0.169 |
| **Diastolic Blood Pressure, mmHg [β coefficient (95% CI)]** |  |  |  |  |  |  |
| Healthiest Behaviors | Reference | - | Reference | - | Reference | - |
| Less Healthy Behaviors | **1.29 (0.66, 1.92)** | **<0.001** | **0.94 (0.21, 1.67)** | **0.012** | 0.56 (-0.15, 1.27) | 0.120 |

*Model 1: unadjusted; Model 2: adjusted for time elapsed between visits, maternal age, education, insurance, income, and perceived racial discrimination; Model 3: additionally adjusted for first trimester blood pressure and body mass index. Bold font indicates significant difference ( $p < 0.05$ ). IRR: incidence rate ratio.*

**Table S6.** Association of incident stage 2 hypertension by behavioral profiles.

| Group | Model 1 | *P* | Model 2 | *P* | Model 3 | *P* |
| --- | --- | --- | --- | --- | --- | --- |
| **Incident Stage Hypertension [IRR (95% CI)]** |  |  |  |  |  |  |
| Healthiest Behaviors | 1.00 (ref) | - | 1.00 (ref) | - | 1.00 (ref) | - |
| Less Healthy Behaviors | 1.25 (0.94, 1.66) | 0.129 | 1.12 (0.79, 1.59) | 0.522 | 0.96 (0.69, 1.35) | 0.818 |

*Model 1: unadjusted; Model 2: adjusted for time elapsed between visits, maternal age, education, insurance, income, and perceived racial discrimination; Model 3: additionally adjusted for first trimester blood pressure and body mass index. Bold font indicates significant difference ( $p < 0.05$ ). IRR: incidence rate ratio.*

**Figure S1.**
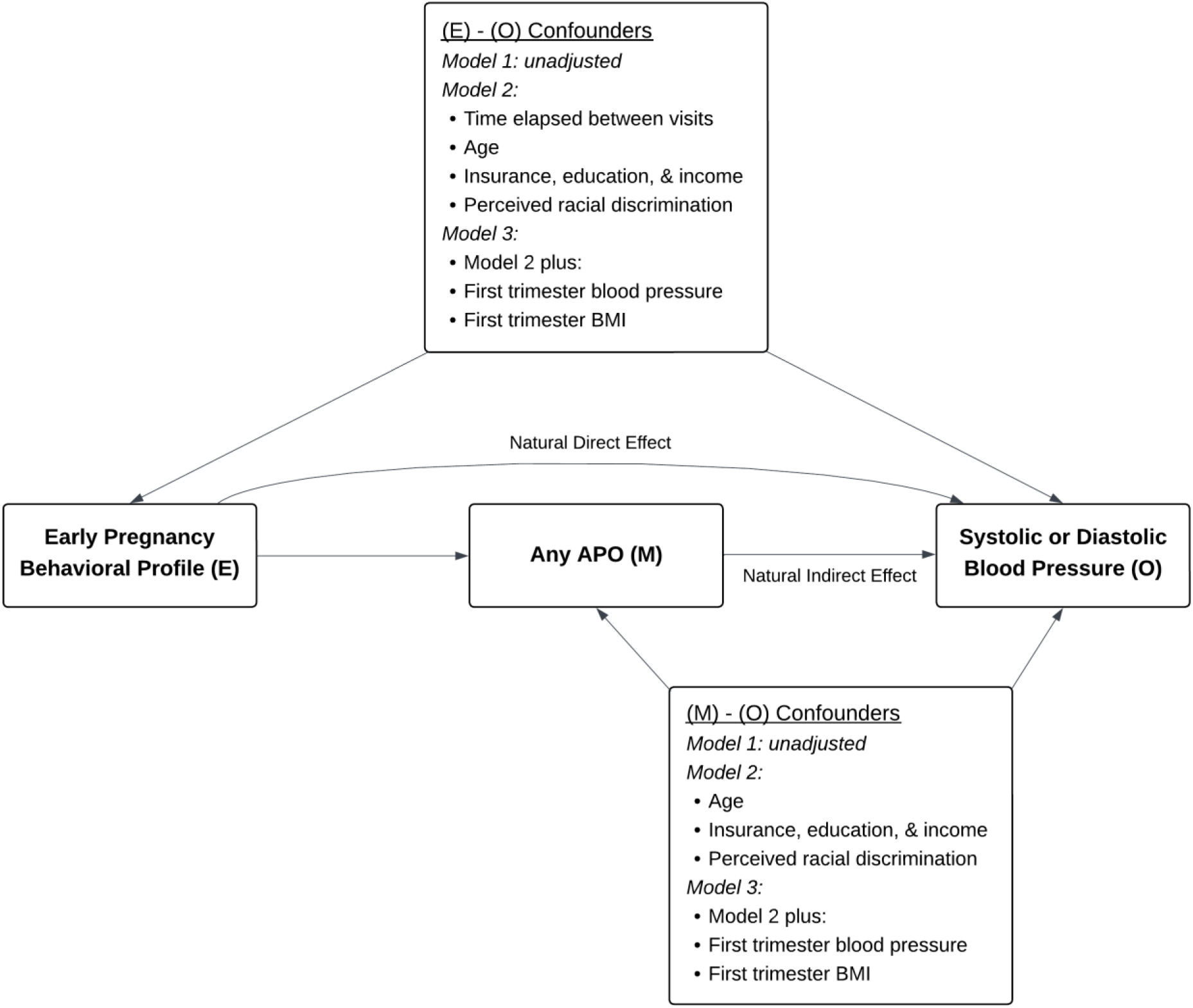
Schematic of mediation analysis to determine the mediating effect of adverse pregnancy outcomes (APO) on the association between early pregnancy behavioral profiles and systolic and diastolic blood pressure 2-7 years following delivery. BMI: body mass index; E: exposure; M: mediator; O: outcome.

